# Proteome-wide Mendelian randomization study implicates inflammaging biomarkers in retinal vasculature, cardiometabolic diseases and longevity

**DOI:** 10.1101/2024.07.13.24310153

**Authors:** Ana Villaplana-Velasco, Nicolas Perrot, Yu Hang, Michael Chong, Emanuele Trucco, Walter Nelson, Jeremy Petch, Hertzel Gerstein, Raina Parminder, Salim Yusuf, Miguel Bernabeu, Albert Tenesa, Konrad Rawlik, Guillaume Pare, Alexander Doney, Erola Pairo-Castineira, Marie Pigeyre

## Abstract

With the increasing proportion of elderly individuals, improving understanding of the biological mechanisms involved in healthy aging is of utmost importance. Retinal vascular complexity obtained from retinal fundus photographs, and evaluated as fractal dimension (ie, *D_f_*), a measure of the geometric pattern of the retinal vasculature, has recently emerged as a valuable indicator of diseases and aging processes. The purpose of our study was to elucidate possible mechanisms underlying those relationships. By integrating retinal *D_f_*with genomic and plasma proteome biomarkers, we uncovered novel pathways involved in determining *D_f_* and its links with cardiometabolic diseases and longevity. First, we performed a genome wide association study (GWAS) of retinal vascular *D_f_* by meta-analysing signals from 74,434 participants from three large epidemiological cohorts, the Canadian Longitudinal Study of Aging (CLSA), the Genetics of Diabetes Audit and Research in Tayside Scotland (GoDARTS), and the UK Biobank (UKBB). We replicated four known loci associated with *D_f_*, and identified one novel locus within or near *DAAM1*, as well as found seven suggestive associations within or near *CSNK1A1L, MAST4, HIVEP1, LOC283038, FNDC3A, SP100,* and *SUGP1* genes. GWAS summary statistics confirmed a negative correlation between *D_f_* and cardiovascular disease, stroke, and inflammation, but a positive correlation with longevity. Next, employing Mendelian randomization which combined genetic determinants of *D_f_*, and *cis*-protein Quantitative Trait Loci from 1,159 circulating proteins from the Prospective Urban and Rural Epidemiological (PURE) cohort (n=12,066), we identified eight causal mediators for *D_f_*, namely IgG Fc receptorIIb, BST1, LILRB2, IL16, MMP12, PON2, ALPP, and PDL2. Notably, MMP12 and IgG Fc receptorIIb have been suggested to connect *D_f_* to cardiometabolic outcomes (i.e., coronary artery disease, stroke, peripheral artery disease, and type 2 diabetes) and longevity, respectively. Pathway enrichment analyses highlighted the role of IgG Fc receptorIIb in immune and inflammatory responses to aging processes, referred to as *inflammaging*. This multi-pronged approach unveils IgG Fc recIIb and MMP12 as key mediators in immune and inflammation pathways, linking lower Df to a higher risk of cardiometabolic diseases and a shorter lifespan. Therefore, retinal *D_f_* may be a convenient way to estimate *inflammaging*, such that lower retinal *D_f_*, may indicate a higher i*nflammaging* status. Targeting MMP12 and IgG Fc ReceptorIIb may promote healthy aging and mitigating the occurrence of cardiometabolic diseases in the elderly population.

**Graphical abstract:** 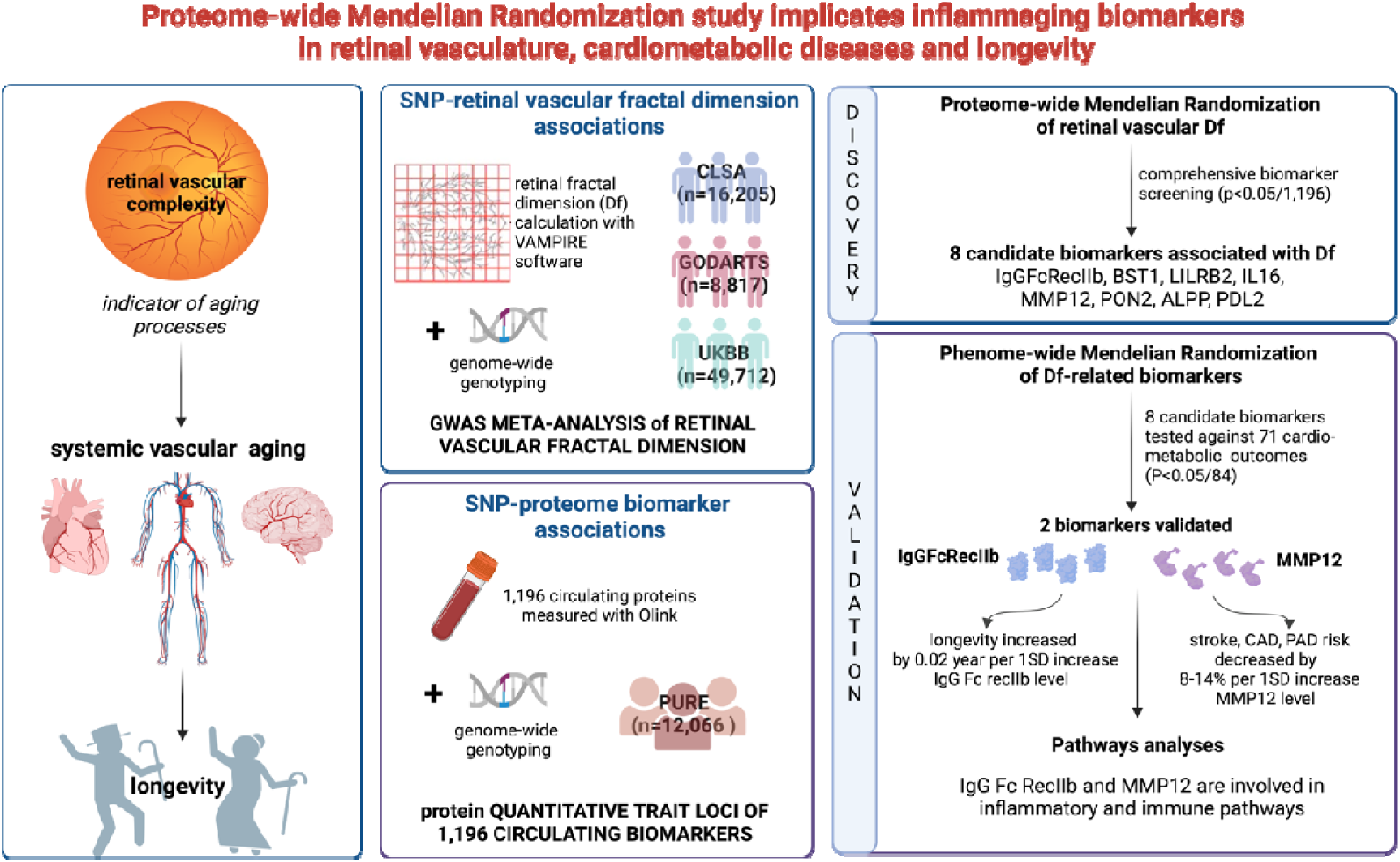

---

Understand the biological pathways that influence healthy aging has become a public health priority ^1^. Aging is associated with an incremental loss of complexity in the dynamics of many physiological systems, rendering them less resilient to environmental stresses ^2^. This loss of resilience progressively increases the vulnerability to late-onset diseases, disability, frailty, and ultimately leads to the death ^3^. Thus, a biomarker signature for “systemic complexity” could potentially provide an indicator of global system resilience.

The retinal vasculature offers a conveniently accessed and imaged perspective on the circulatory system of the human body, providing valuable insights not only into the vascular health of the retina itself but also of other organs ^4^. The automated quantification of density and overall complexity of the retinal vasculature from retinal photographs—achieved through the measurement of its vascular fractal dimension (abbreviated as *D_f_*), with a higher value reflecting a more complex branching pattern—offers a powerful tool in this regard ^5^. Early evidence suggested that aging-related changes in the retinal vasculature result in a decrease of the *D_f_* measures ^6^. More recently, research has provided compelling evidence that reduced retinal vascular *D_f_* is a risk factor for various age-related diseases, including hypertension, type 2 diabetes, coronary artery disease, stroke, and cognitive decline ^5,7^. Those findings underscore the clinical significance of assessing retinal vasculature complexity through its *D_f_*, as a valuable indicator of aging-related vascular changes. However, the biomolecular mechanisms underlying the relationship between retinal *D_f_* and the occurrence of age-related diseases remain largely unknown.

We explored the innovative concept of employing *D_f_* as an indicator of aging, enabling the causal biomolecular links between aging and cardiometabolic health to be explored. We aimed to uncover novel pathways involved in aging processes by combining *D_f_* measures with high-throughput genetic and plasma proteome biomarkers. Specifically, we leveraged genetic factors influencing *D_f_*to explore the causal connections between microvascular complexity, circulating protein levels, and cardiometabolic outcomes and longevity using Mendelian randomization (MR) ^8^.

In our comprehensive multimodal approach, we conducted a large genome-wide association study (GWAS) meta-analysis on retinal *D_f_* from three expansive epidemiological cohorts, namely the Canadian Longitudinal Study of Ageing (CLSA), the Genetics of Diabetes Audit and Research Tayside Study (GoDARTS), and the UK Biobank (UKBB). We next performed a proteome-wide Mendelian randomization (MR) analysis, which combined genetic determinants of *D_f_,* with greater than one thousand circulating protein biomarkers from individuals enrolled in the Prospective Urban and Rural Epidemiological (PURE) study. This approach allowed us to pinpoint candidate biomarkers for *D_f_*, and investigated whether such biomarkers are also involved in the links between *D_f_*and cardiometabolic diseases and longevity. Then, we conducted pathway enrichment analyses to provide a deeper understanding of the interactions in which the identified biomarkers are involved within the systemic molecular network (Figure 1). Collectively, our study indicated critical pathways and mechanisms, and provided insights to promote healthy aging and longevity. This study also informed on new therapeutic targets to mitigate cardiometabolic diseases.

**Figure 1:**
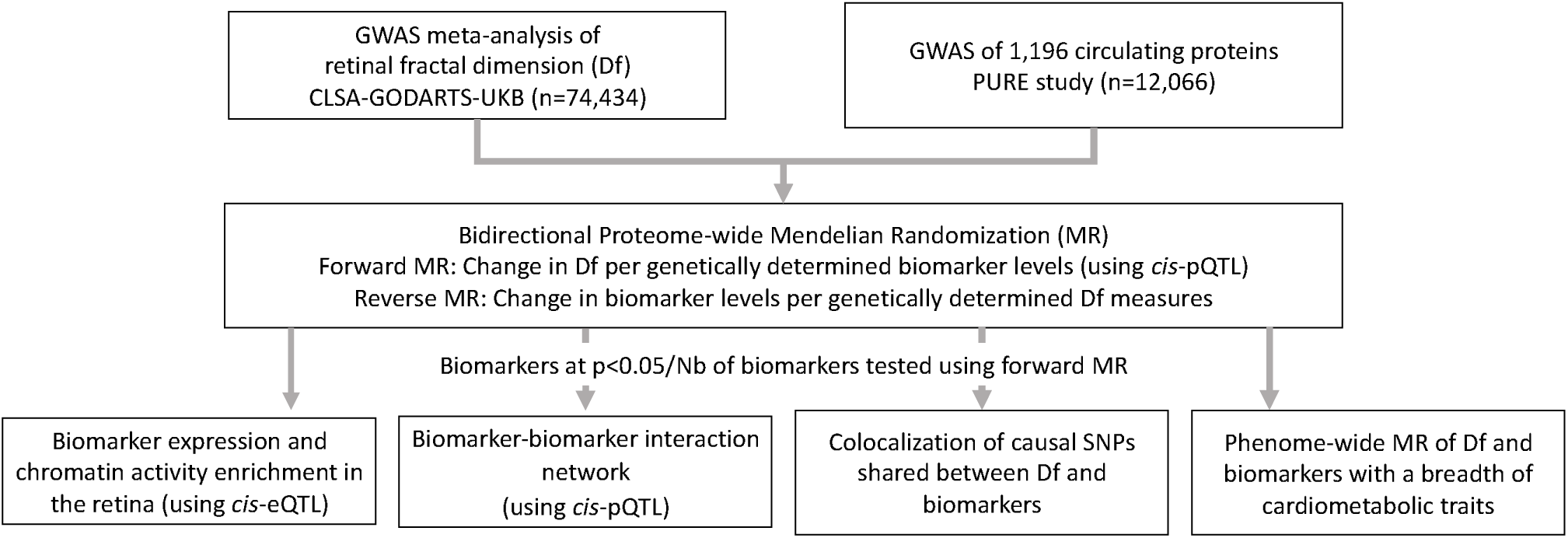
Study design

## Results

### Genome-wide association study meta-analysis identified eight novel loci associated with retinal fractal dimension

We performed GWAS meta-analysis for retinal *D_f_* from 74,434 individuals of European ancestry who participated in CLSA, GoDARTS or UKBB cohorts. The retinal fundus images available from all individuals in those cohorts were processed by the automatic VAMPIRE software to compute *D_f_*, as previously described elsewhere ^9^. The demographic characteristics of each cohort are summarized in Suppl Tables 1-3. First, we conducted the GWAS by fitting a linear model with polygenic effect adjustment in each cohort separately. We then performed an inverse-variance weighted meta-analysis of the GWAS results. The quantile-quantile plot showed an adequate control of genomic inflation λ*_GC_* = 1.065 (Suppl Figure 1). The meta-analysis revealed five independent genome-wide significant associations (at P-Value < 10^-8^) (Figure 2, Table 1), including one novel locus and four replicated loci reported in previous GWASs of retinal *D_f_* ^5,10^. The novel association corresponded to an intronic variant located on chromosome 14 (rs2295848, P-Value = 1.41 x 10^-9^) in the *DAAM1* gene, which has been previously related to brain imaging phenotypes ^11,12,13,14^. Two of the replicated loci were close to *HERC2* (rs12913832, P-Value = 2.34 x 10^-59^) and *OCA2* (rs35717941, P-Value = 1.20 x 10^-11^), two genes on chromosome 15 that has previously been associated with retinal *D_f_*, pigmentation, and several eye conditions, including cataract, visual acuity or retinal vessel tortuosity ^14,15,16^ Additionally, two other genetic variants near *SLC45A2* on chromosome 5 (5:33956560, P-Value = 7.58 x 10^-9^) and *SLC12A9* on chromosome 7 (rs80308281, P-Value = 1.51 x 10^-10^) were associated with retinal *D_f_*, both genes being related to hair pigmentation ^17,18^. The *SLC12A9* lead variant has also been reported to be associated with arterial blood pressure and resting heart rate ^19^.

**Figure 2:**
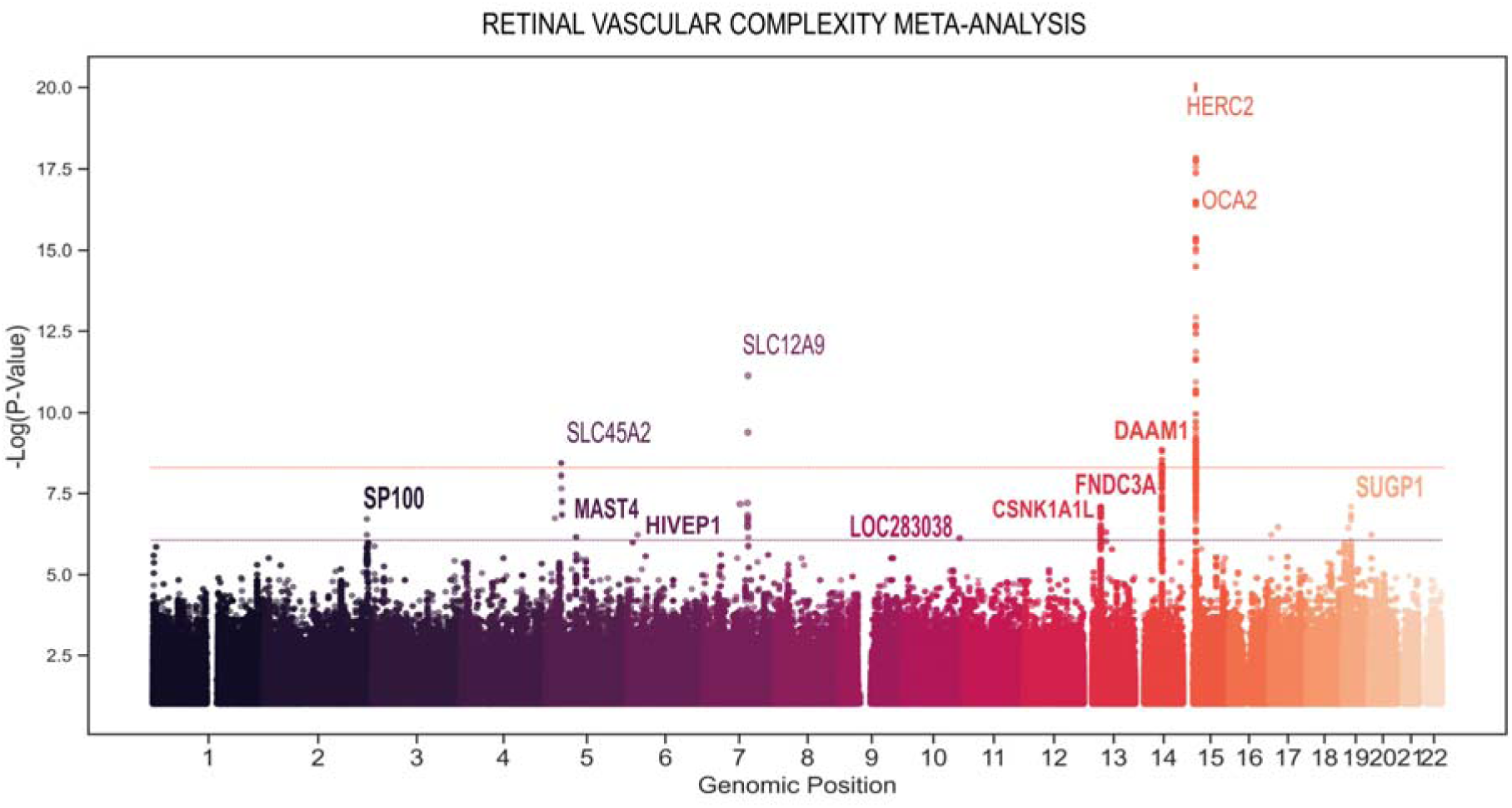
Manhattan Plot of the GWAS meta-analysis (CLSA-GODARTS-UKB) for *D_f_*. Points are truncated at –log10(P)=20 for clarity. Bold gene names indicate novel significant and suggestive associations.

**Table 1.**
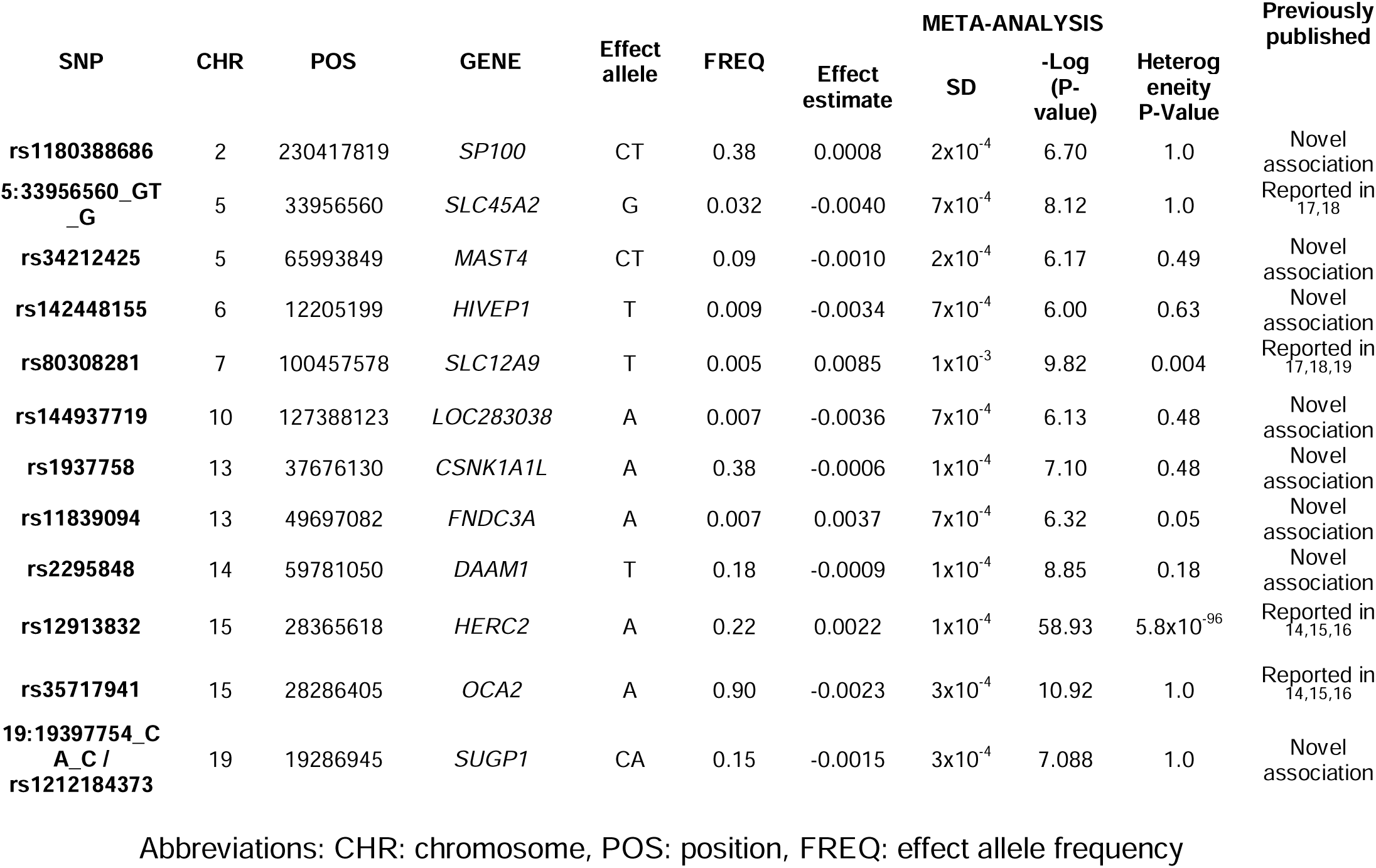
GWAS meta-analysis results

In addition to these genomic regions, we found seven suggestive associations (at P-Value < 10^-6^), including an intergenic variant near *CSNK1A1L* located on chromosome 13 (rs1937758, P-Value = 7.94 x 10^-8^); an insertion at *MAST4* located on chromosome 5 (rs34212425, P-Value = 6.76 x 10^-7^), which has previously been associated with hypertension and coronary artery disease ^20,21,22^; a locus in a regulatory region on chromosome 6 (rs142448155, P-Value = 1.00 x 10^-6^) that is close to *HIVEP1,* which is a gene related to inflammation and immunity ^23^; a variant in *LOC283038* on chromosome 10 (rs144937719, P-Value = 7.41 x 10^-8^); a variant in *FNDC3A,* located on chromosome 13 (rs11839094, P-Value = 4.78 x 10^-7^), which has been associated with chronic autoimmune inflammatory diseases ^24^; a variant at *SP100* on chromosome 2 (rs1180388, P-Value = 1.99 x 10^-7^), for which the gene may play a role in angiogenesis and viral infection ^25,26^; and a variant in *SUGP1* on chromosome 19 (rs1212184373, P-Value = 2.39 x 10^-7^), a gene associated with cholesterol concentration and coronary artery disease ^27,28^. No significant heterogeneity was found between cohorts, except for the variants located in the *HERC2/OCA2* gene (Heterogeneity P-value = 5.79 x 10^-96^). Those loci were significantly associated with *D_f_* in the UKBB and CLSA, but with a stronger effect in the UKBB (Suppl Figure 2).

Subsequent gene-level analysis of GWAS using the Multi-marker Analysis of GenoMic Annotation (MAGMA)^29^ tool, showed an enrichment of significant genes in pathways related to melanin biosynthesis and inflammatory response (Suppl. Tables 4,5). Finally, we estimated the genetic correlations of Df with systemic inflammation and cardiometabolic outcomes. We found nominally significant negative correlations of *D_f_* with C-reactive protein (r_g_ = -0.13 ± 0.05), atherosclerosis (r_g_ = -0.24 ± 0.03), coronary artery disease (r_g_ = -0.12 ± 0.06), stroke (r_g_ = -0.23 ± 0.09), and a positive correlation with longevity (r_g_ = 0.12 ± 0.06) (Suppl Table 6). Such findings were consistent with the findings that we previously reported for analysis conducted in a subset of UKBB individuals ^10^.

### Mendelian Randomization and colocalization demonstrate causal associations for immune and inflammatory biomarkers with retinal fractal dimension

We screened a panel of 1,159 plasma biomarkers involved in inflammation and cardiometabolic pathways ^30^ for putative causal associations with *D_f_* using a two-sample bidirectional MR analysis. A subset of 264 biomarkers had suitable genetic instruments to be retained in the forward MR, including circulating biomarkers as exposures, and retinal *D_f_* as outcome. We found evidence of an association between genetically-predicted biomarker concentrations and *D_f_* for eight of those biomarkers beyond the Bonferroni-corrected significance threshold (at P-Value < 0.05/264). Genetically-determined circulating levels of IgG Fc recIIb, BST1, LILRB2, IL16, MMP12, and PON2 were positively associated with *D_f_*, while genetically-determined circulating levels of ALPP and PDL2 were negatively associated with *D_f_* (Figure 3A, Suppl Table 7). When stratifying analyses according to diabetes status, associations for LILRB2, MMP12, PON2, and ALPP were consistent, while additional positive associations for TCN2 levels, and negative associations for MATN3, TMPRSS5, and UNC5C with *D_f_* were observed in individuals without diabetes (P-Value < 0.05/136) (Suppl Table 8). Positive associations of FCN2, SERPINA9, DDC, IL2RA, and LGMN, and negative associations of C1QA, NCAM, and CRTAM levels were also observed with *D_f_* in individuals with diabetes (P-Value < 0.05/244) (Suppl Table 9).

**Figure 3:**
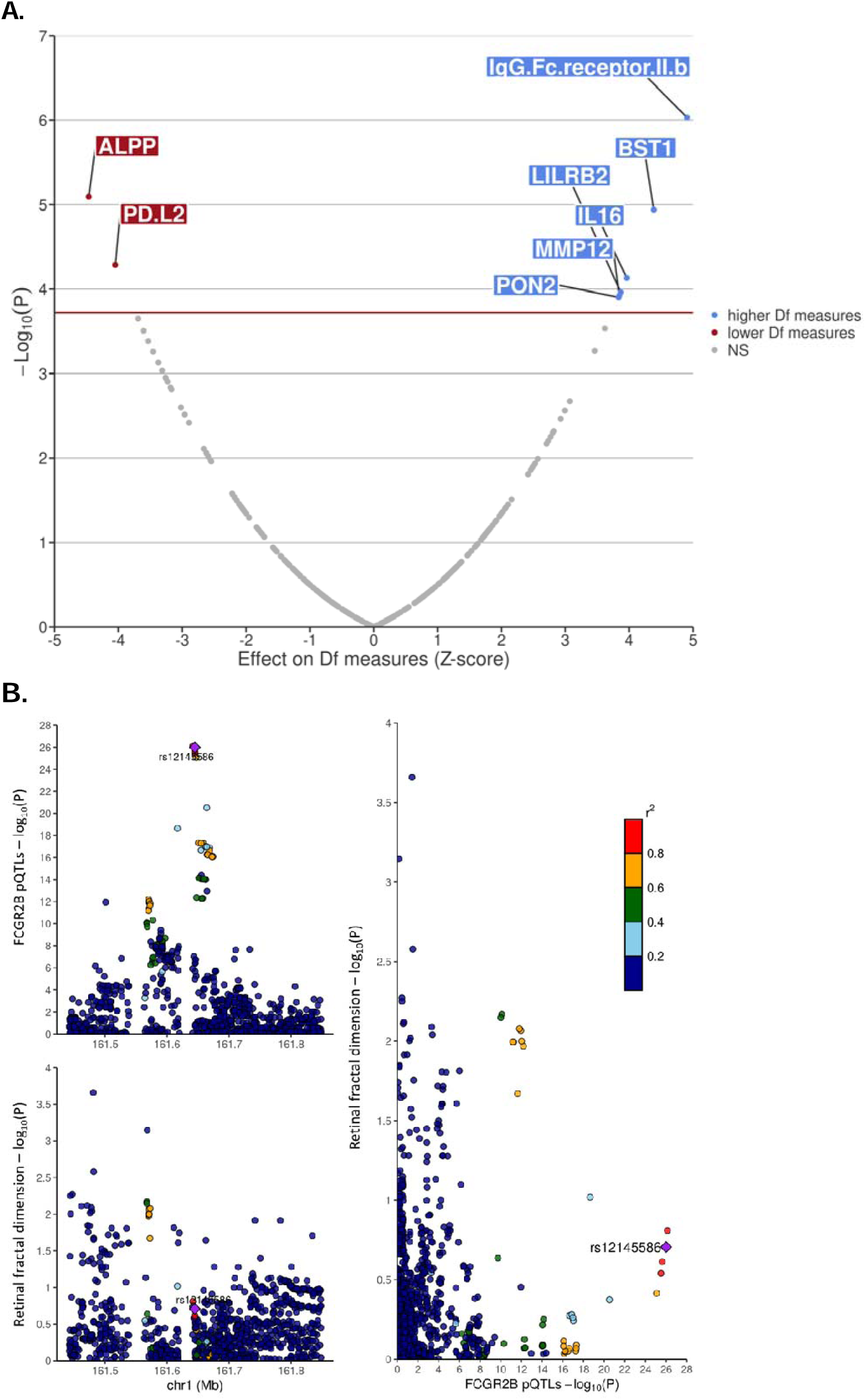
MR and colocalization of biomarkers associated with *D_f_*. A. Volcano plot of circulating biomarkers associated with Df, identified by MR. B. Colocalization of FCGR2B with Df.

Using a total of 1,159 unique biomarkers, a reverse MR analysis, using *D_f_*as exposure and biomarkers as outcomes, was used to rule out a reverse causation of *D_f_* on circulating biomarker levels. Although, this analysis did not reveal significant associations between genetically-determined *D_f_* measures and biomarkers (P-Value < 0.05/1,159 = < 4.3 ×10^-5^), the most significant association was a negative relationship between *D_f_*and FOXO3, such that FOXO3 circulating levels decreased by 12.27 (+/-3.21) SD per 1 SD increase in genetically-determined *D_f_* (P-Value = 1.35×10^-4^) (Suppl Table 10). This is noteworthy given that FOXO3 has an established role in longevity ^31^ and vascular aging ^32^.

### Phenome-wide Mendelian Randomization revealed that three biomarkers associated with retinal fractal dimension are also shared with cardiometabolic outcomes and longevity

We conducted a phenome-wide MR to investigate the associations of retinal *D_f_* and biomarkers related to *D_f_*, with a broad range of cardiometabolic outcomes, encompassing 71 phenotypes for which the GWAS summary statistics were extracted from publicly available consortium data. First, phenome-wide MR of *D_f_* on cardiometabolic outcomes showed significant inverse relationships with estimated glomerular filtration rate (P-Value < 0.05/71) (Suppl Table 11). Next, among the eight *D_f_*-related biomarkers, three were also significantly associated with cardiometabolic disease phenotypes (P-value < 0.05/(8*71)). Higher genetically-predicted IgG Fc recIIb levels were associated with longer parental lifespan (mean change of 0.02 (+/-0.003) year per 1 SD increase in biomarker level; P-Value = 5.5 x 10^-7^) and decreased LDL-Cholesterol levels (mean change of -0.01 (+/-0.002) mmol/L per 1 SD increase in biomarker level; P-Value=1.24 x 10^-5^). Higher MMP12 levels were associated with a lower risk of stroke (OR per 1 SD increase in biomarker level, 0.92; 95%CI 0.90-0.95; P-Value = 5.96 x 10^-11^), and in particular, with stroke of ischemic cause (OR, 0.91; 95%CI 0.89-0.94); P-Value = 6.90 x 10^-9^), as well as with a lower risk of peripheral artery disease (OR, 0.86; 95%CI 0.83-0.90; P-Value = 1.55 x 10^-13^), type 2 diabetes (OR, 0.95; 95%CI 0.93-0.97; P-Value = 1.65 x 10^-5^), and coronary artery disease (OR, 0.92; 95%CI 0.87-0.96; P-Value = 1.28×10^-4^). Higher LILRB2 levels were associated with lower HDL-cholesterol levels (mean change of -0.06, +/-0.008 mmol/L; P-Value = 6.43 x 10^-13^) and higher triglycerides levels (0.02, +/-0.004 mmol/L; P-Value = 3.14 x 10^-7^) (Figure 4; Suppl Table 12).

**Figure 4:**
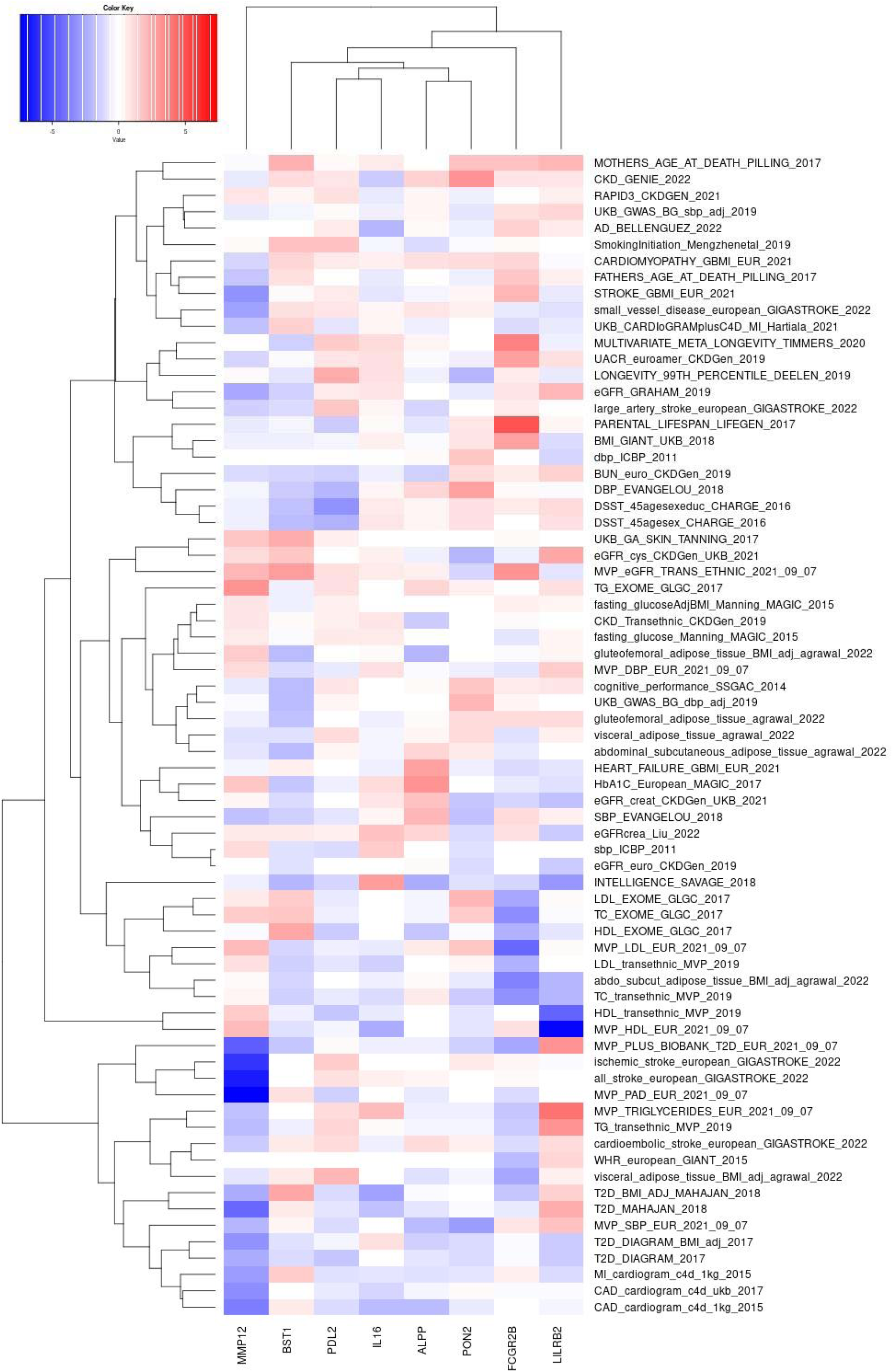
Heatmap of the associations between significant biomarkers for *D_f_* and cardiometabolic disease outcomes.

Multivariate MR investigating the effects of the identified biomarkers on diseases, adjusted for their effects on *D_f_*, confirmed that the effects of IgG Fc recIIb on longevity were partially mediated by an effect on *D_f_*, with an attenuation of the effect of IgG Fc recIIb on longevity by 34% when adjusting on *D_f_* (*D_f_*adjusted mean change of 0.01 (+/-0.002) year per 1 SD increase in biomarker level; P-Value = 3.31 x 10^-12^). The effects of MMP12 levels on cardiometabolic outcomes were also partially mediated by its effects on *D_f_*, but to a lesser extent (ranging from 9% to 15% of effect attenuation after adjustment on *D_f_*) (Suppl Table 13).

Moreover, colocalization analysis confirmed that circulating IgG Fc recIIb levels (encoded by *FCGR2B* gene) and retinal *D_f_* shared a common causal genetic variant with a posterior probability superior to 80% (i.e., H3+H4≥0.8) (Figure 3B, Suppl Table 14). To investigate which cells might drive this *FCGR2B* protein Quantitative Trait Locus (pQTL) association, we conducted further colocalization analyses between *FCGR2B* pQTL and single-cell expression quantitative trait loci (sc-eQTL) in peripheral blood mononuclear cells (PMBCs)^33^. This supported the presence of the causal variant effect in *FCGR2B* across different cell types, and most of which were involved in the immune response (Suppl Table 15, Suppl Figure 3).

### Network analysis of biomarkers confirms enrichment for immune and inflammatory pathways

We reinforced the MR findings with protein-protein interaction networks, which revealed significant enrichment for pathways involved in immune and inflammatory responses, such as cytokine-cytokine receptor interaction pathway (False Discovery Rate (FDR) = 4.12×10^-5^), and immune-related disease (FDR = 0.015) (Figure 5, Suppl Tables 16,17). We also investigated the expression levels and chromatin activity of the *D_f_* -associated biomarkers in the retinal tissue. Out of the eight MR significant biomarkers, three (IgG Fc recIIb, PON2, and IL16) were expressed in retinal tissue and associated with open chromatin regions and related to immune and metabolic networks (Suppl Table 18) ^34^.

**Figure 5:**
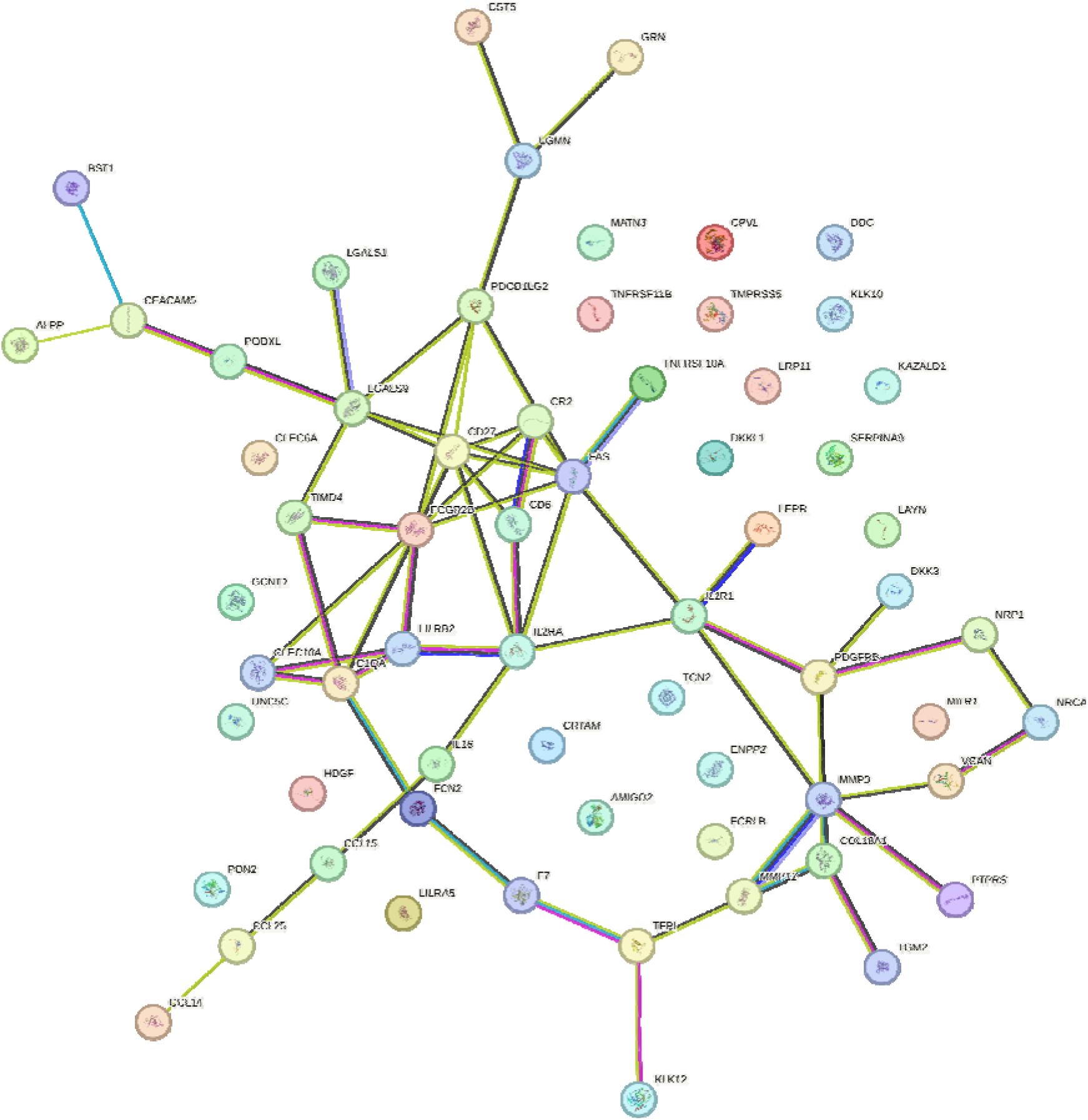
Network analysis of associated biomarkers (P-value <0.001) from MR results using the UKB and CLSA cohorts.

## Discussion

To the best of our knowledge, this study is the largest GWAS meta-analysis for retinal *D_f_* to date. We identified eight novel loci associated with *D_f_*, and replicated the associations of four previously identified variants ^5,10^. We also confirmed genetic correlations between *D_f_* and systemic inflammation, cardiovascular disease, stroke and longevity, and identified circulating MMP12 and IgG Fc recIIb levels as key mediators linking *D_f_* to cardiometabolic and longevity outcomes. Notably, enrichment analyses also highlighted the role of IgG Fc recIIb in immune and inflammatory responses to aging, processes referred to as *inflammaging*. Finally, reverse MR suggested that circulating FOXO3 levels may be a blood biomarker for changes in retinal vascular *D_f_*.

The findings from the *D_f_* GWAS meta-analysis were consistent with previous genetic analyses on retinal *D_f_*, thus, supporting the biologically coherent relationships of *D_f_* - associated variants with various ocular and cardiovascular traits and diseases ^5,10,35^. Interestingly, we found that several loci were also associated with either cardiovascular risk factors (e.g., hypertension, and dyslipidaemia) or diseases (e.g., coronary artery diseases), as well as inflammatory and immune responses20,21,22,23,24,25,26,27,28.

Previous studies focused on the links between *D_f_* and diseases, and showed that a lower *D_f_* was associated with a higher risk of mortality, incident hypertension, congestive heart failure, renal failure, type 2 diabetes, sleep apnea, anemia, and multiple ocular conditions, and was causally associated with skin cancer and retinal detachment ^5,10^. However, none of those studies investigated the molecular mechanisms linking *D_f_* to such diseases. Our study is the first MR analysis investigating the relationships between the proteome and *D_f_*, and provides novel evidence on the role of eight circulating biomarkers with changes in *D_f_*, including IgG Fc recIIb, BST1, LILRB2, IL16, MMP12, PON2, ALPP, and PDL2. Additionally, we investigated the associations between those eight biomarkers and health-related traits. Notably, IgG Fc recIIb and MMP12 also emerged as causal mediators of diseases, linking *D_f_* to longevity and to various chronic diseases, including stroke, peripheral artery disease, and type 2 diabetes, respectively. The IgG Fc recIIb effect size on lifespan was in the range of previously reported biomarkers for longevity ^36^. The MR findings were reinforced by single-cell colocalization analysis in PMBCs, which confirmed the putative causal role of IgG Fc RecIIb on *D_f_* changes driven by its expression in immune cells; and by enrichment analysis that indicated that IgG Fc RecIIb expression was associated with different regions of the retinal regulatory network.

Biologically, IgG Fc RecIIb is a densely expressed receptor that plays a role in the immune response and antiviral activity of malt cells, basophils, eosinophils, macrophages, monocytes, dendritic cells, and B cells in the spleen and lymph nodes ^37^. Previous studies showed that variants leading to impaired FcγRecIIb activity cause the accumulation of IgGs, B cell apoptosis, and a pro-inflammatory states ^38,39^. Efforts to pharmacologically modulate FcγRecIIb either directly by regulating immune responses or indirectly by influencing FcγRecIIb interactions through other pathways, using monoclonal antibodies and protein fusion approaches, have produced drugs, such as Obinutuzumab or Rituximab with applications in immune diseases, and hematologic cancers ^40,41^.

The MR findings were consistent with previous plasma proteome studies that showed that circulating MMP12 levels were linked to cardiovascular outcomes, including heart disease, peripheral artery disease, and stroke ^42,43,44,45^. Higher genetically-predicted levels of MMP12 have consistently been associated with a lower risk of stroke, and it has been suggested as potential therapeutic target ^46^. Biologically, MMP12 belongs to a family of endopeptidases that degrade structural components of the extracellular matrix ^47^. MMP12 plays key pro- and anti-inflammatory roles in the body through the cleavage of cytokines and subsequent regulation of cell migration and signaling, and may contribute to atherosclerotic cardiovascular disease pathogenesis through the regulation of macrophage migration, plaque development, and rupture ^47^.

Chronic inflammation related to aging has been characterized as a clinical condition, named *inflammaging*, associated with a high risk of prematurely developing age-related diseases and adverse health outcomes ^48,49^. This hypothesis agreed with our complementary biomarker analysis that revealed associations between IgG Fc recIIb and lifespan, and MMP12 levels and cardiovascular diseases. Considering that genetic predictions of IgG Fc recIIb and MMP12 levels are also associated with *D_f_*, we propose *D_f_* as a potential accessible imaging marker of the inflammatory status related to aging, such that a lower *D_f_* would be indicative of a higher inflammaging status.

Moreover, reverse MR suggested that genetic variants increasing *D_f_* were negatively associated with circulating FOXO3 levels, suggesting that circulating FOXO3 levels might reflect microvascular homeostasis. This is consistent with previous research on FOXO3 that reported its association with human longevity ^31^, and its critical role in vascular aging ^32^. FOXO3 is required for maintaining vascular homeostasis under stressful conditions and preventing vascular aging through the regulation of oxidative resistance, apoptosis, autophagy, energy metabolism, and extracellular matrix remodeling processes by targeting the expression of effector genes. Under normal conditions, FOXO3 is inactive due to its negative regulation by the IIS-PI3K-Akt pathway. When cells are exposed to stress, including growth factor deprivation, metabolic stress, and oxidative stress, FOXO3 translocates into the nucleus and exhibits increased transcriptional activity. Dysregulation of FOXO3 contributes to atherosclerosis, vascular calcification, hypertension, and vascular aging-related heart diseases, kidney diseases, and cerebrovascular diseases. Thus, FOXO3 has been considered as a promising therapeutic target for age-related vascular diseases^50^.

Even though this was the largest genetic study on retinal vascular traits to date, to validate our results and extrapolate them to a valid clinical outcome, it is of utmost importance that new studies include individuals of non-European ancestry. Although we measured 1,159 selected circulating proteins, this still represents a small subset of the ∼20,000 proteins in the human proteome ^51^. From a methodological perspective, MR relies on several assumptions that cannot all be assessed, although our analyses generally showed concordant effect estimates between different MR approaches. For instance, when assessing the significance of the Egger intercept test, we employed a conservative approach of not correcting for multiple testing; however, our power to detect directional pleiotropy may have been insufficient ^52^. If undetected directional pleiotropy did not affect the results, the lack of concordance between the MR-Egger estimates and the other MR estimates might be attributable to a violation of an assumption of MR-Egger ^53^ . We may also have limited power to assess co-localization between variants associated with protein level and *D_f_*, leading to the possibility that significant MR findings might reflect the presence of separate causal variants in linkage disequilibrium with one another ^54^. We measured protein expression levels in plasma, but not in the retina. However, the use of *cis-*pQTLs, which are likely to be shared across tissues ^55^, as instrumental variables in these MR analyses, supports the possibility that the MR associations reflect the actions of the proteins of interest in the retina. Additionally, there are little publicly available data for the retinal transcriptome and proteome to perform further enrichment analysis. This lack of available information and rarity of studies is driven by the invasiveness of collecting retinal tissue, and for the complex extraction of proteins and DNA from such samples.

In conclusion, our study identified IgG Fc recIIb and MMP12 as key mediators in immune and inflammation pathways, linking lower Df to a higher risk of cardiometabolic diseases and a shorter lifespan. Therefore, retinal *D_f_*may be a convenient way to estimate *inflammaging*, such that lower retinal Df may indicate a higher inflammaging status. Finally, these findings pave the way for new strategies targeting IgG Fc recIIb and MMP12 to promote healthy aging by mitigating age-related immune and inflammatory pathways involved in cardiometabolic diseases and longevity.

## Online methods

### Study populations

The UK Biobank (https://www.ukbiobank.ac.uk) is a large multi-site cohort study that consists of 502,655 individuals aged between 40 and 69 years at baseline, and recruited from 22 centres across the UK between 2006-2010. The study was approved by the National Research Ethics Committee, reference 11/NW/0382, and informed consent was obtained from all participants as part of the recruitment and assessment process. From these, a baseline questionnaire, physical measurements, and biological samples were undertaken for each participant. Ophthalmic examination was not included in the original baseline assessment and was introduced as an enhancement in six UKBB centres across the UK. This examination consisted in capturing paired retinal fundus with a 45° primary field of view obtained using a Topcon 3D OCT-1000 MKII (Topcon Corporation). These analyses were completed using fundus images collected at the first and the second ophthalmic examination visits from 49,712 participants of white European ancestry with at least one eye image of good quality. The data were processed as published in our previous analysis ^10^.

Participants from Genetics of Diabetes Audit and Research in Tayside Scotland (GoDARTS) ^56^, Genetics of Scottish Health Research Register (GoSHARE) ^57^ were used for GWAS analysis. GoDARTS is a cohort study in the Tayside region of Scotland that began recruiting participants in 1996 and continued to 2015. The study included 10,149 individuals with type 2 diabetes and 8,157 control subjects without type 2 diabetes at the time of recruitment ^56,58^. GoSHARE Data on clinical and lifestyle parameters were collected at the time of recruitment, and participants also provided consent to their electronic health record linkage. Retinal photographs used for the Scottish national diabetes retina screening (DRS) are available for all patients with diabetes in GoDARTS. DRS photographs were obtained using a standardized protocol with a 45° view centered on the macula. Participants also provided a sample of blood for genome-wide genotyping that was performed using multiple separate genotyping arrays, including the Affymetrix version 6.0, Illumina OmniExpress BeadChips, Illumina Infinium Broad BeadChips, Illumina Human OmniExpressExome-8 version 1.0 BeadChip, and Illumina OmniExpressExome-8 version 1.2 BeadChip. Genetic data were available for 7,722 T2D participants after quality control. GWAS data were then obtained using the Affymetrix Genome-Wide Human SNP Array 6.0 the Illumina HumanOmniExpress and Broad. The Affymetrix GWAS chip contains 932,979 single nucleotide polymorphisms (SNPs), and the Illumina GWAS chip contains 731,296 SNPs. Then imputation of additional and missing genotypes were performed by SHAPEIT ^59^ and IMPUTE2 ^60^ using the 1000 Genomes reference panel ^61^. In addition, 707 Type 2 diabetes cases and 3,478 controls were genotyped using custom genotyping arrays from Illumina. Such arrays include the Immunochip, Cardio-Metabochip (Metabochip), and Human Exome array. The Immunochip contains 196,524 genetic markers from loci that have previously been associated with at least one of 13 autoimmune diseases, including Type 1 diabetes ^62^, while the Metabochip contains 196□725 SNPs from loci that have prior evidence of associations with T2D, coronary artery disease/myocardial infarction, and 21 related traits ^63^. The Human Exome Array contains 247,870 genetic markers from across the exome, allowing for studies to focus on identifying protein-altering variants.^64^ The genotyping process for the GoDARTS and GoSHARE cohorts involved various platforms, including Affymetrix 6.0, Illumina Omni Express-12VI, and GSA v2.0.

After applying quality control criteria, a total of 7,722 participants (6,249 from GoDARTS and 1,473 from GoSHARE) were considered for analysis. The quality control criteria used to exclude individuals from analysis were: Individual genotype call rate < 95%; discrepancy in gender; heterozygosity > 3 standard deviations from the mean; and highly related samples identified through identity by descent analysis. SNP-level quality control was performed by excluding markers with a call rate of less than 95% and those with a Hardy-Weinberg P-Value < 1×10^-6^. PLINK versions 1.7 and 1.9 were employed for quality control assessment and data preprocessing for imputation. Ancestry outliers were detected using principal component analysis in each cohort. The genotype data from all three cohorts were then imputed against the HRC r1.1 reference panel. Monomorphic markers and those with an imputation quality score below 0.4 were excluded from the post-imputation data ^16^.

The Canadian Longitudinal Study on Aging (CLSA) is a large, national, stratified, random sample of 50,000 Canadians aged 45 to 85 years at the time of recruitment (2010-2015) and followed until 2033 (or until death). The CLSA aims to investigate the associations between risk factors and the incidence of chronic diseases ^65^. A subset of 30,000 participants (i.e., comprehensive subset) had physical examinations and biological specimen collection, including fundus photographs (1 for each eye) obtained using the Topcon TRC-NW8 non-mydriatic retinal camera. A total of 50,957 retinal photographs from 25,717 CLSA participants, were analysed using VAMPIRE (Vascular Assessment and Measurement Platform for Images of the Retina, version 3.1, University of Edinburgh and University of Dundee, U.K.) to compute image quality (good/moderate/poor) and the fractal dimension (Df) of the retinal vascular pattern. Participants with poor image quality for both eyes were excluded from subsequent analyses. Among the comprehensive subset, 26,622 CLSA participants (with 93% of Europeans) were successfully genotyped using the UK Biobank Array ^66^. The quality control steps have been detailed elsewhere ^67^. Briefly, phasing and imputation were conducted using the TOPMed reference panel at the University of Michigan Imputation Service. We used the TOPMed reference panel version r2, and then pre-phased and imputed the genotype data using EAGLE2 and Minimac, respectively, for both autosomal and X chromosomes. Samples with low call rates (<95%), sex mismatches, or cryptic relatedness were removed. Imputed SNPs were excluded based on low call rates (<95%), deviation from Hardy-Weinberg (p<10-6), low minor allele frequency (MAF<0.0001), and low imputation quality (Rsq < 0.6).

The Prospective Urban Rural Epidemiology (PURE) study is a large prospective study of individuals in 27 low-income, middle-income, and high-income countries. Participant recruitment and selection has been described in detail elsewhere ^68^. Briefly, the PURE study collected socio-demographics, physical examination, routine laboratory results (e.g., lipid profile), lifestyles, depression and stress questionnaires, disease status and age at diagnosis, medication usage, and family health history at recruitment and each follow-up visit. A biobanking initiative was developed for a subset of PURE participants recruited between January 5, 2005, and December 31, 2006 to assess genomic and proteomic markers of chronic disease risk, based on a case-cohort design. Blood samples from participants were transported from 14 countries (i.e., Argentina, Bangladesh, Brazil, Canada, Chile, Colombia, Iran, Pakistan, Philippines, South Africa, Sweden, Tanzania, United Arab Emirates, and Zimbabwe) to the Population Health Research Institute (Hamilton, ON, Canada) and stored at −165°C. Samples were considered eligible if they belonged to individuals from the major self-reported ethnicity in the residing country (e.g., European ancestry in Sweden). Samples were deemed ineligible if they were unsuitable for analysis or were non-fasting. Hereby, we selected a random sample from the pool of 55,246 eligible participants, and then included all individuals who had incident events of interest that were not selected as part of the random sample. The final sample set included 12,066 PURE participants. The research ethics committees at each study location, including Hamilton Health Sciences, approved the project (Hamilton, ON, Canada). Written informed consent was obtained from each study subject.

### Fractal dimension

Retinal fractal dimension (commonly abbreviated as *D_f_*) is a measure of vasculature density and branching complexity, as previously described elsewhere ^69^. *D_f_* measurements were obtained after fundus image segmentation with VAMPIRE V3.1 software for each cohort, using validated methods for image pre-processing, vessel segmentation map and *D_f_* estimation ^9,70,71^. The three datasets included participants with both eyes analysed. In the cases of CLSA and UKB, if both images were of a similar good quality, the mean Df was used; otherwise, the image with the highest quality was chosen. For GoDARTS, we included the mean Df if both images were available.

### Fundus image quality

Fundus image quality measurement in CLSA was defined through manual evaluation of the vessel segmentation map generated by the VAMPIRE V3.1 software. As for the UKB, this quality evaluation, named image quality score (IQS), was derived from a prior study ^72^. The software perform automatically detects the retinal vasculature, creating a binary vessel map for each image, thus, following the pipeline described in ^10^. Images with IQS >0 were classified as good quality and were included in the pipeline. When participants had paired good quality images, the mean Df and the mean image quality score were used for subsequent analysis in the CLSA and UKB datasets. GoDARTs did not include image quality measurements, but retinal images were subject to a manual quality control step during the ophthalmic examination to ensure suitability for phenotype.

### GWAS for Fractal Dimension

We established a pipeline to complete the same GWAS across each cohort. The GWAS model was an additive mixed linear model that adjusted for age, sex, fundus image quality score, and the first 10 genetic principal components. Because GoDARTS is a cohort to study diabetes, the GoDARTS GWAS was also adjusted for the presence of diabetic retinopathy. This process was completed using REGENIE software ^73^, using genotyped SNP data for the first step, and imputed SNP information for the second step.

### Meta-analysis

Variants included for the meta-analysis were selected independently in each cohort. Variants were autosomal SNPs present in the genotyping and imputing panel with a Hardy-Weinberg Equilibrium (HWE) >10^−6^, minor allele frequency (MAF) >5×10^−3^, call rate >0.9, and imputation score >0.8. The number of total SNPs analysed after quality control was 30,275,286 SNPs in the UKB; 11,950,600 SNPs in the CLSA; and 8,059,472 SNPs in the GoDARTS.

The meta-analysis was completed using a fixed effect inverse variance-based model using METAL software ^74^. Additionally, we completed a random-effects inverse variance meta-analysis in the significantly heterogenic genetic variants from the fixed-effects meta-analysis using the meta package in R ^75^. We visualised the effect of SNPs across cohorts using forest plots in the meta package in R ^75^.

Loci in the meta-analysis were defined using the clump function of PLINK1.9, and UK Biobank European ancestry genotypes as the linkage disequilibrium (LD) reference panel. Clumping parameters were: r2 = 0.1, P = 1×10^-6^, P2 = 0.01, and kilobase (KB) = 10,000 bp. Associated genes for independent loci were established using the nearest gene strategy. Gene level analyses were conducted by MAGMA software ^76^.

### Genetic correlation

Genomic inflation and genome-wide genetic correlation estimates (rg) for completed GWASs were estimated using the LD Score and HDL software ^77,78^.

### Phenome-wide association studies

Phenome-wide association analysis (PheWAS) was completed to investigate the effects of significant SNPs from the meta-analysis on other traits. To this end, independent lead SNPs were searched in GWAS Catalog ^79^ and GeneATLAS ^80^, databases that contain numerous GWAS summary statistics for diverse traits and diseases. Genetic variants must have a P-value less than 1×10^-8^ on the trait to assume a significant association.

### Circulating protein biomarker measurement

A 1.8 mL aliquot of plasma from each PURE participant was processed in the Clinical Research Laboratory and Biobank in Hamilton, ON, Canada. Plasma protein concentrations of 1,159 unique circulating biomarkers were measured using an immunoassay based on proximity extension assay (PEA™) technology (Olink^®^ PEA panels; Uppsala, Sweden, 13 panels of 96 biomarkers each, including CVDII, CVDIII, Metabolism, Inflammation, OncologyII, Cardiometabolic, Organ Damage, Development, Cell Regulation, Immune response, Neurology, OncologyIII, and Neuro Exploratory). The data generated are expressed as relative quantification on the log2 scale of normalized protein expression (NPX™) values. Although NPX values are relative quantification units, the Olink PEA platform has been extensively validated and previous work shows strong relationships between measurements from the multiplex Olink panel and singleplex assays of the same markers with absolute units ^82^. Individual samples were excluded on the basis of quality controls for immunoassay and detection, as well as the degree of hemolysis ^30^. NPX values were rank-based normal transformed for further analyses.

### Circulating protein level genome-wide association study of 1,159 unique biomarkers

PURE participants suitable for proteomics analyses were directly genotyped for > 800,000 polymorphisms on the Thermofisher Axiom Precision Medicine Research Array (release 3). The genotyping quality control has been described elsewhere ^30^. Briefly, sample-level quality control checks included assessments of sample completeness (call rate > 0.95), potential sample mix-ups (discrepancies between reported vs. genetically determined sex and/or ethnicity), genetic duplicates, and sample contamination (excess heterozygosity). Samples exhibiting non-ambiguous discrepancies between genetic and self-reported ancestry were removed. Variant-level quality control checks included assessments of variant completeness (call rate > 0.985), plate and batch effects, non-Mendelian segregation within families (Mendelian errors), Hardy Weinberg Equilibrium (HWE) deviations (HWE P-value < 1×10^-5^), and variant frequency (minor allele frequency > 0.001). After sample and variant quality control procedures, up to 3,514 samples from Europeans and 749,783 variants remained. The average genotyping call rate among passing samples was 0.996535. Analyses were restricted to participants from European, Latin or Persian ancestry as linkage disequilibrium differs between ethnic groups, and thus, might prevent MR assumption violation. We conducted a genome-wide association study analysis (also known as protein Quantitative Trait Loci (pQTLs)), testing the association between each of the 1,159 plasma protein concentration and each of the common genetic variants in 9,150 PURE participants of European, Latin or Persian ancestry, using a linear regression model adjusted for age, sex, and 20 ancestry-specific principal components.

### Bidirectional Mendelian randomization and colocalization analyses

To construct the genetic instruments in forward MR, we used *cis*-protein Quantitative Trait Loci (*cis*-pQTL) variations that were associated with circulating protein levels (P-Value < 5×10^-6^), were located up to 200 kb downstream or upstream of the gene that codes for each measured protein, and were independent (pairwise r2 < 0.1). For reverse MR, we used SNPs associated with Df with a P-Value <5×10^-8^. SNPs were pruned for linkage disequilibrium at a threshold of r2 < 0.1. A total of 35 to 41 SNPs (F-statistics: 31.0 to 31.8) were retained.

As a genetic instrument linked to a protein-altering variant can influence the measurement of the protein binding affinity, genetic variations in the pleiotropic major histocompatibility complex locus, missense and splicing site variants, or variants in linkage disequilibrium with those variants (r2 ≥ 0.9) were excluded in both forward and reverse MR analyses.

For both the forward and reverse MR, we applied the inverse variance weighted IVW method as the primary method. The MR robust adjusted profile scoring (RAPS) method was also performed. A Bonferroni threshold of significance was employed for the MR analysis. MR analyses have been performed using TwoSampleMR and mr.raps R packages.

MR was performed separately in three ethnic groups, European, Persian and Latin, and the results were meta-analyzed using the fixed effect method from the metafor R package rma.uni function. Multivariable Mendelian randomization is a statistical method used to investigate the causal relationship between multiple exposures or risk factors and an outcome of interest ^53^. In multivariable Mendelian randomization, the aim is to assess the mediating role of intermediate variables in the causal pathway between an exposure and an outcome. To conduct multivariable Mendelian randomization analysis, genetic variants that are associated with both the exposure and/or the intermediate outcomes were used with the MVMR R package ^83^. Phenome-wide MR used similar methods to those used in forward MR, with genetic instruments being the *cis*-pQTLs. Genetic associations for outcomes were extracted from publicly available summary statistics of genetic consortia (Suppl. Tables 11, 12).

To determine if the causal variant underlying a significant GWAS association for Df is shared with the biomarker pQTL, colocalization analysis was performed using the Pair-Wise Conditional analysis and Colocalization analysis (PWCoCo) pipeline ^84^. We excluded the presence of colocalization when the posterior probability that an association existed between a genetic variant and both Df and biomarker concentration (H3+H4) was less than 80% ^84^. We complemented the colocalization with an additional analysis between pQTL biomarkers and blood single-cell eQTLs from peripheral blood mononuclear cells (PMBCs) generated by the Onek1k resource ^33^. This step was completed following the Coloc pipeline, which is very similar to that of PWCoCo. Essentially, we extracted those SNPs that colocalized in the GWAS-pQTL analysis, which were then used for an additional colocalization between sc-eQTLs and pQTLs. Causal variants colocalized in expression and proteomic levels if the posterior probability of sharing the causal signal PPH4 > 0.8.

### Expression and chromatin activity in retinal tissue

To further support the role of associated Df loci and *cis*-pQTL biomarkers in retinal tissue, we assessed the evidence of their expression levels and the activity of that genomic region using the Gene Regulatory Networks in Human Retina (Retina GRN)^85^ and human retinal networks, and enriched annotation from eyeIntegration software ^86^. Both software programs combine published gene expression data, expression quantitative trait loci (eQTL) information, and chromatin accessibility and loops data, in the case of Retina GRN, from human retinal tissue to derive the regulatory networks.

### Protein-protein interaction network analysis of MR biomarkers

We evaluated the biological mechanisms and the interactions between associated MR biomarkers through a protein-protein interaction network study and an enrichment analysis. We used biomarkers that had a P-Value < 0.0001 for each MR analysis in European, European-Latin, or European-Latin-Persian populations to investigate the general processes. STRING ^87^ and DAVID ^88^ software were then used to identify the protein-protein interactions of MR biomarkers and their genetic and pathway enrichment. Enriched pathways and functional annotations were considered if FDR and Bonferroni correction were < 0.01.

## Data availability

The GWAS meta-analysis results can be download from XXX (DATASHARE, University of Edinburgh) upon publication. The UK Biobank Df calculations can be accessed upon request to UK Biobank resources.

PURE data contain the personal health information of participants from 27 high-, middle-, and low-income countries collected over 20 years. Consent for public disclosure of this information was not obtained at the time of data collection and can no longer be practically collected. Deidentified individual-level genotype and proteomic data for the PURE cohort cannot be shared publicly or through a controlled data repository, but the authors welcome inquiries for collaboration.

## Data Availability

All data produced in the present study will be available upon publication

## Acknowledgements

This research has been conducted using the UK Biobank Resource under project 788. This research was funded in part by the Medical Research Council grant (MR/N013166/1) to A.V.V. M.O.B. gratefully acknowledges funding from: Foundation Leducq Transatlantic Network of Excellence (17 CVD 03); EPSRC grant no. EP/X025705/1; British Heart Foundation and The Alan Turing Institute Cardiovascular Data Science Award (C-10180357); Diabetes UK (20/0006221); Fight for Sight (5137/5138); the SCONe projects funded by Chief Scientist Office, Edinburgh & Lothians Health Foundation, Sight Scotland, the Royal College of Surgeons of Edinburgh, the RS Macdonald Charitable Trust, and Fight For Sight; the Neurii initiative which is a partnership among Eisai Co., Ltd, Gates Ventures, LifeArc and HDR UK.

This research has been conducted using the CLSA dataset 1906019_McMaster_GPare_Baseline (Baseline Comprehensive (COM) version 4.2), under Application Number 1906019. The CLSA is led by Drs. Parminder Raina, Christina Wolfson and Susan Kirkland. M.P. was supported by the E.J. Moran Campbell Internal Career Research Award from McMaster University, and the Early Career Research Award from Hamilton Health Sciences (HHS). Retinal images analyses of the CLSA were supported by a New Investigator Fund from HHS (NIF-18453 to M.P.), and the Canadian Institutes of Health Research (CIHR) (ACD-170312 and PJT-178302 to M.P.).

The PURE study is an investigator-initiated study funded by the Canadian Institutes of Health Research (CIHR), Heart and Stroke Foundation of Ontario, the Ontario Ministry of Health and Long-Term Care, and through unrestricted grants from industry including AstraZeneca, Sanofi-Aventis, Boehringer Ingelheim, Servier, and GalaxoSmithKline. The biomarker project was led by PURE investigators at the Population Health Research Institute (Hamilton, Canada) in collaboration with Bayer scientists. Bayer directly compensated the Population Health Research Institute for measurement of the biomarker panels, and scientific, methodological, and statistical work. Genetic analyses were supported by CIHR (G-18-0022359) and the Heart and Stroke Foundation of Canada (399497) in the form of funding to GP.

## Author contributions

Conceptualization: E.P.C., M.P.

Methodology: A.V.V, N.P., M.C, E.T., J.P., G.P., A.D., E.P.C., M.P.

Software/formal analysis: A.V.V, N.P., Y.H., M.C, E.T., W.N., E.P.C., M.P.

Visualization: A.V.V, N.P., E.P.C., M.P.

Funding acquisition: G.P., M.P. Supervision: G.P., A.D., E.P.C., M.P.

Writing of the original draft: A.V.V, N.P., E.P.C., M.P.

Writing, review and editing: Y.H., M.C., E.T., W.N., J.P., H.G., R.P., S.Y., M.B., A.T., K.R., G.P., A.D.

The authors had full control, and funders played no role in the design, analysis, or interpretation of this work.

All authors have reviewed and approved the final version of the manuscript.

